# Population ageing, incarceration and the growing digital divide: understanding the effects of digital literacy inequity experienced by older people leaving prison

**DOI:** 10.1101/2023.04.13.23288514

**Authors:** Ye In (Jane) Hwang, Amanuel Hagos, Adrienne Withall, Stephen Hampton, Phillip Snoyman, Tony Butler

## Abstract

**Background:** Digital inequity refers to the inequality and exclusion experienced by those who lack the same opportunities or circumstances to support the development of digital skills as the rest of modern society. One rapidly growing and highly vulnerable group to digital inequity is older people attempting to reintegrate into society after release from prison, where technology access is limited. Inadequate support for digital skills in this population entails widespread consequences for public health, human rights, social welfare and recidivism. This qualitative study is the first to: examine digital inequity experienced by older people who have been incarcerated, understand the effects of this on reintegration to society, and begin informing appropriate solutions.

**Method:** Semi-structured interviews were conducted with N=15 older people (mean age= 57) who had been released from an Australian prison in the last two years, regarding their experiences of digital literacy since leaving prison. Reflexive thematic analysis was conducted under a critical realist lens.

**Results:** The analysis resulted in six themes that illustrated the extent of digital inequity experienced by this population, and key challenges for improving digital literacy: ‘surviving in a digital world’, ‘stranger in a foreign world’, ‘questioning the digital divide’, ‘overcoming your “old” self’, ‘don’t like what you don’t know’, and ‘seeking versus finding help’.

**Conclusions:** The digital inequity that older people experience during and after incarceration creates additional challenges for a growing group who are already medically and socially marginalised. Prioritisation of this group for digital literacy initiatives both during incarceration and in the community will have benefits for their health, social and financial reintegration. Their unique life experiences should be considered in designing and delivering these programs.

## Introduction

### A growing digital divide

Digital literacy is a vital skill in modern societies where technology is widely and increasingly used to perform essential functions such as seeking information, communicating with others, and accessing services. In this research, ‘digital literacy’ refers broadly to the having the skills to navigate information and communication technologies (henceforth referred to as ‘technology’), which include a broad range of hardware, software and telecommunications such as the internet and smartphones.

In contrast to the rest of modern society, prisons are unique, secure environments that typically limit their occupants’ access and use of technology. For this reason, there has been increased recognition of prisons as places that can cause digital inequity or exclusion (e.g., McDougall et al., 2017; Reisdorf & Jewkes, 2016). Often referred to as a ‘digital divide’, people who are incarcerated experience a period of disconnection from the digital world and its advancements (Kerr & Willis, 2018). Resultantly, prisoners have been referred to as ‘one of the most impoverished groups in the digital age’ (Reisdorf & Jewkes, 2016, p. 1). This issue is gaining the attention of researchers, governments and advocates worldwide, with studies from countries such as Finland, the United Kingdom, Indonesia, and Australia referring to such a divide (Ariani, 2021; Järveläinen & Rantanen, 2021; Office of the Inspector of Custodial Services WA, 2018; Reisdorf & Jewkes, 2016).

The digital inequity experienced during imprisonment has important effects for re-entry to society after leaving prison due to its impact on factors such as health inequity, social participation and economic reintegration (Azzopardi-Muscat & Sørensen, 2019; Champion & Edgar, 2013). With 30 million people entering and leaving prisons globally each year, these are a large and growing population who are already at risk of significant social and medical marginalization in the community (World Health Organization, 2021). Successful reintegration is already a challenge in many countries. A systematic review of 2-year reconviction rates globally, reported figures between 20-63% for reconviction and 14-45% for reimprisonment (Yukhnenko et al., 2019). Among the poorest performing countries in this review was Australia, who most recently reported that 45% of people returned to prison within 2 years of their release (Productivity Commission, 2022). Researchers and policymakers have increasingly acknowledged the likely impact of digital literacy on successful post-release life. Digital literacy can enhance reintegration by encouraging prosocial and productive use of time and family connections (see Kerr & Willis, 2018). A systematic review of 29 such publications published since 2015 concluded that digital literacy (referred to as “digital rehabilitation”) “makes re-entry successful and guarantees enhanced post-prison life in a digitalised society” (Zivanai & Mahlangu, 2022, p. 2).

### Older prison leavers face significant digital inequity

One group disproportionately affected by the digital divide in prisons is older people. Older prisoners are among the fastest-growing subpopulations of prisoners worldwide, with significant health, social and economic implications (Ginnivan et al., 2021; Maschi et al., 2013; Psick et al., 2017). In the past five years, countries such as Italy, the UK and Australia have reported that between 18-25% of their prison population are older, and this continues to grow for reasons including general population ageing, a trend toward harsher sentencing practices and the rise of historical convictions (Ginnivan et al., 2018; Lorito et al., 2018; Luallen & Cutler, 2017; Roth, 2014).

Research from the general population shows that older people are already at a general disadvantage in terms of digital literacy, and prone to digital exclusion. Older people are less likely to access or use technology such as smartphones and the internet (Choudrie et al., 2020; Hunsaker & Hargittai, 2018) and perform poorly on measures of digital inclusion (Thomas et al., 2021). A recent systematic review identified that older age is a consistent predictor of digital division (Lythreatis et al., 2022).

Older people in prison are a unique and high-needs group for whom intersectionality of old age and imprisonment introduces additional challenges for reintegration (Scaggs & Bales, 2017; Skarupski et al., 2018; Solares et al., 2020). First, social isolation is a commonly discussed consequence of digital illiteracy (Lythreatis et al., 2022). The heightened risk of social exclusion for prison leavers of all ages with poor digital literacy has already been recognized by researchers (Jewkes & Reisdorf, 2016). This is further challenging for older people, who often experience disconnection from their social networks both whilst in prison and after release (Hayes et al., 2013; Wyse, 2018). Release in older age also tends to be associated with longer sentences, further deepening social disconnection and a longer period of exclusion from digital advancements (Lorito et al., 2018). One qualitative study of released older men in the US found disconnection from community was particularly difficult, with serious implications for recidivism (Lares & Montgomery, 2020).

Older prisoners also have heightened and complex health needs. Older prisoners have significantly higher rates of chronic conditions compared to both younger prisoners and people of similar age in the community (Australian Institute of Health and Welfare, 2019; Prost et al., 2021; Solares et al., 2020). Studies have also shown high rates of emergency department use in the 6 months after leaving prison for those over 55 (Humphreys et al., 2018). Given their high and acute health needs, the shift towards electronic health systems, and online means of accessing health services and information in many countries creates an additional barrier for this group where digital literacy may not be adequate.

Finally, as with many organisations following the rise of the COVID-19 pandemic, there is an increased trend towards using technology in corrective services such as fulfilling parole requirements and to deliver rehabilitation programs both in prison and after release (Kerr & Willis, 2018). This leaves those with inadequate digital literacy at a crucial disadvantage that may impact return to prison.

### Knowledge gaps

To date, there has been no focused investigation of the technology use and digital literacy challenges of older people after they are released from prison. However, existing and related studies have highlighted the importance of specifically examining this age group after release. For example, Lares and Montgomery (2020) identified “Technology challenges” as a subtheme in their qualitative study of N=19 older people who had left prison. In another qualitative study by Hagos et al. (2021), Australian correctional staff identified lack of digital literacy as a key barrier for health and social service access in older people leaving prison. One cross-sectional survey of N=255 adult prisoners in Finland regarding their use of digital health care and social welfare services found that age was an additional difficulty for digital inclusion, and concluded that digital skills initiatives are particularly warranted for older and longer-term prisoners (Rantanen et al., 2021).

There has been limited examination of the digital literacy of prisoners in Australia with most being reviews and discussions regarding increasing technology use in prisons. One discussion paper published in the Australian Institute of Criminology recognized the rising challenge embracing technology in prisons as ‘now one of the key issues facing correctional managers’ (Kerr & Willis, 2018, p. 1) and provided a comprehensive review of the challenges and opportunities that this has brought about in prisons both in Australia and internationally. Among their conclusions they state that improved access to technology for prisoners is a key priority to enable rehabilitation and reintegration after release. In the same year, the Office of the Inspector of Custodial Services in Western Australia also published a report regarding the digital divide in prisons (Office of the Inspector of Custodial Services WA, 2018). Its focus was mostly on availability and access issues relating to technology in prisons, including six recommendations relating to wider implementation and evaluation of technology in prisons.

Whilst some of the likely challenges faced by older prison leavers can be gleaned from existing studies, we can expect that the unique sociocultural and age-related contexts that surround older prison leavers will interact with digital literacy in unique ways. Specific, evidence-based interventions are needed. There is a clear and urgent need to understand the digital literacy and technology use situation and challenges specifically for older people leaving prison, and to develop nuanced understandings of how this affects re-entry to the community.

### Aims

This exploratory qualitative study aimed to develop a contextualized and rich understanding of the key issues in digital literacy, specifically for older people leaving prison. Specific objectives of the current study were to:

1. Identify factors that affect digital literacy in older prison leavers
2. Understand the experience and effects of digital literacy on post-release life

The age cutoff for this study was 50 years old, or 45 years old for Aboriginal and/or Torres Strait Islander people. Whilst lower than traditionally considered in studies of ‘older’ adults, this is in line with the general consensus for what is considered ‘older’ in the prison context, due to their various and compounded health and medical vulnerabilities (Australian Institute of Health and Welfare, 2019; Merkt et al., 2020).

## Methods

### Ethics

This study was granted ethical approval from: The University of New South Wales Human Research Ethics Committee [HC220042], Corrective Services NSW Ethics Committee [D2022/0294030], and the Justice Health and Forensic Mental Health Network of NSW Ethics Committee [G477/22].

### Recruitment and sampling

This study sought to interview older adults who had left prison in Australia. Inclusion criteria:

1. Released from an Australian correctional centre in the past 24 months
2. Aged 50 years or older at release (45+ if an Aboriginal or Torres Strait Islander person)
3. Spent at least 12 months incarcerated (sentenced or on remand)
4. English ability sufficient to participate in a 60-minute phone interview

We employed purposive sampling to identify and recruit eligible participants via two pathways. The first included contacting organisations that provide services to individuals who may have experiences of imprisonment. These included specific post-prison transition support services, community housing organisations, and other relevant non-government-organisations across Australia. Study information and Participant Information Statement and Consent Forms (PISCFs) were sent via email to the organisations. These were passed on by staff to potential participants, who were advised to contact the research team directly via phone or email to express interest. Organisations were also encouraged to share the study invitation and PISCF via their websites and social media platforms.

Second, assistance was provided by Community Corrections offices run by Corrective Services NSW, who are responsible for supervising offenders in the community. Staff at these offices across the state were sent study information and PISCFs and asked to introduce the study to potential participants by showing them the study flyer and giving them a copy of the PISCF to read and keep. Interested participants contacted the research team directly via email or phone call to express interest.

Upon receiving expressions of interest, the research team screened participants according to inclusion criteria. Once deemed eligible, the research team went through the PISCF again and answered any further questions. If the participant was willing, the contact details of the participant were recorded, and an interview date arranged. Verbal consent was recorded prior to commencing the interview.

### Data collection

One-on-one, semi-structured interviews were conducted via telephone by two researchers. Interview questions were open ended and focused on participants’ experiences of using technology after leaving prison, and how it has affected their life after prison. Interviews were audio-recorded on handheld devices. Participants were reimbursed $75AUD via direct bank transfer for completing the interview. Two of the authors (JH and AH) were involved in interviewing participants, and therefore were potentially able to identify individual participants from the data that was collected. The other authors did not have access to identifying information for individual participants.

### Analysis

All interview recordings were transcribed by the interviewers and imported to NVIVO 12 software for analyses. Reflexive thematic analysis was selected for analysis. Thematic analysis is an analysis method that seeks to extract ‘patterns of meaning’ in qualitative data via the production themes (Braun & Clarke, 2019, p128). Reflexive thematic analysis specifically seeks to uncover common themes in the data across a set of participants whilst emphasising a contextualized understanding of the data where the researcher’s interpretations play a central role in the process (Braun & Clarke, 2021a). It favours a more interrogative and interpretative stance towards the data than other forms of thematic analysis (see Braun & Clarke, 2021b).

An inductive approach was chosen, such that analysis was conducted without trying to fit the themes into any pre-existing theory. Notwithstanding, no qualitative work can be conducted in a ‘theoretical vacuum’ (Braun et al., 2020), and the researchers acknowledge the influence of intersectionality as a perspective that may affect the analysis and interpretation to some extent. Briefly, intersectionality refers to the idea that people belong to multiple social categories that are based on both individual factors and social structures/systems, and these intersect to create unique circumstances and power dynamics that must be considered in interpreting a person’s experience (Atewologun, 2018). In terms of ontology and epistemology, analysis and interpretation of the data rested on critical realism, assuming that there are independent ‘realities’ in the world, but such reality is accessed imperfectly through observation and consideration of the contexts and experiences of the person.

Braun and Clarke’s 20-item checklist for evaluating thematic analyses was used to ensure quality in the present study. As is recommended in reflexive thematic analysis, the first author conducted the analyses alone. Analysis occurred in five steps.

1. Data familiarization: The entire dataset was read alongside the recordings. With the research aims in mind, notes were made regarding impressions of the data.
2. Coding: Transcripts were imported to NVIVO software. Line by line, meaningful sections of text were selected and assigned a descriptive label. The labels were a mix of semantic (i.e., describing the text as is) and latent (i.e., some level of interpretation of the implicit meaning behind the text) levels of meaning.
3. Initial theme generation: Codes across the dataset that appeared to centralise on a similar concept were organized into initial themes. Themes needed to include codes from across several participants to be considered a ‘common pattern of meaning’.
4. Developing and reviewing themes: The initial themes were reviewed and refined in an iterative process with the aim of creating broader themes. Themes were considered in light of the research question, how much data was available for each, how homogenous or internally coherent each theme was, and how distinct each theme was from the others.
5. Refining themes: Each theme was given a name and brief definition to describe its content. Quotes were selected to represent each theme. These themes were then organized visually and structurally using a thematic map.

## Results

### Participants

Interviews were conducted between May-August 2022. N=15 older people participated in interviews (see Table 1) which lasted approximately 30-60 minutes each.

**Table 1.**
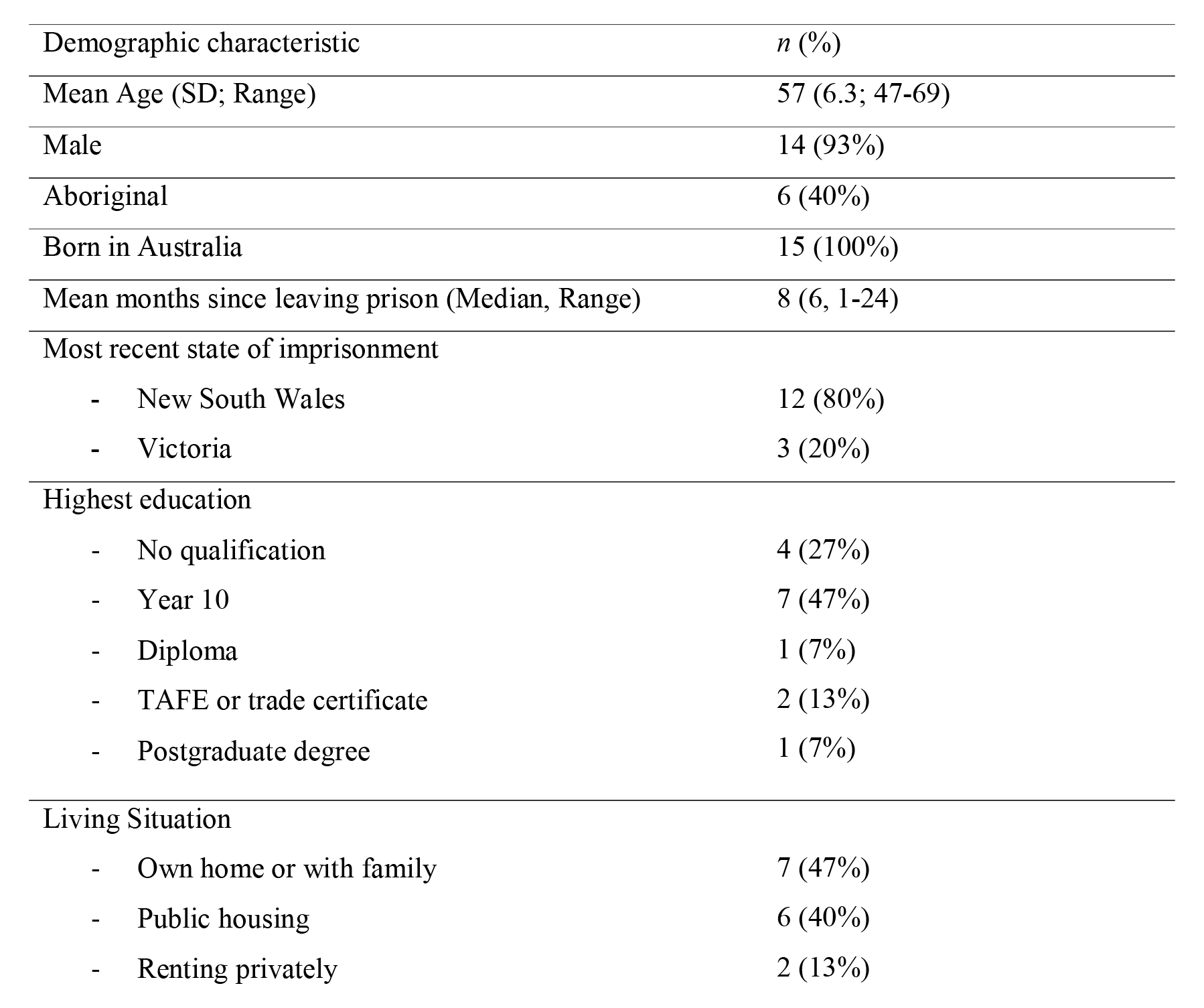
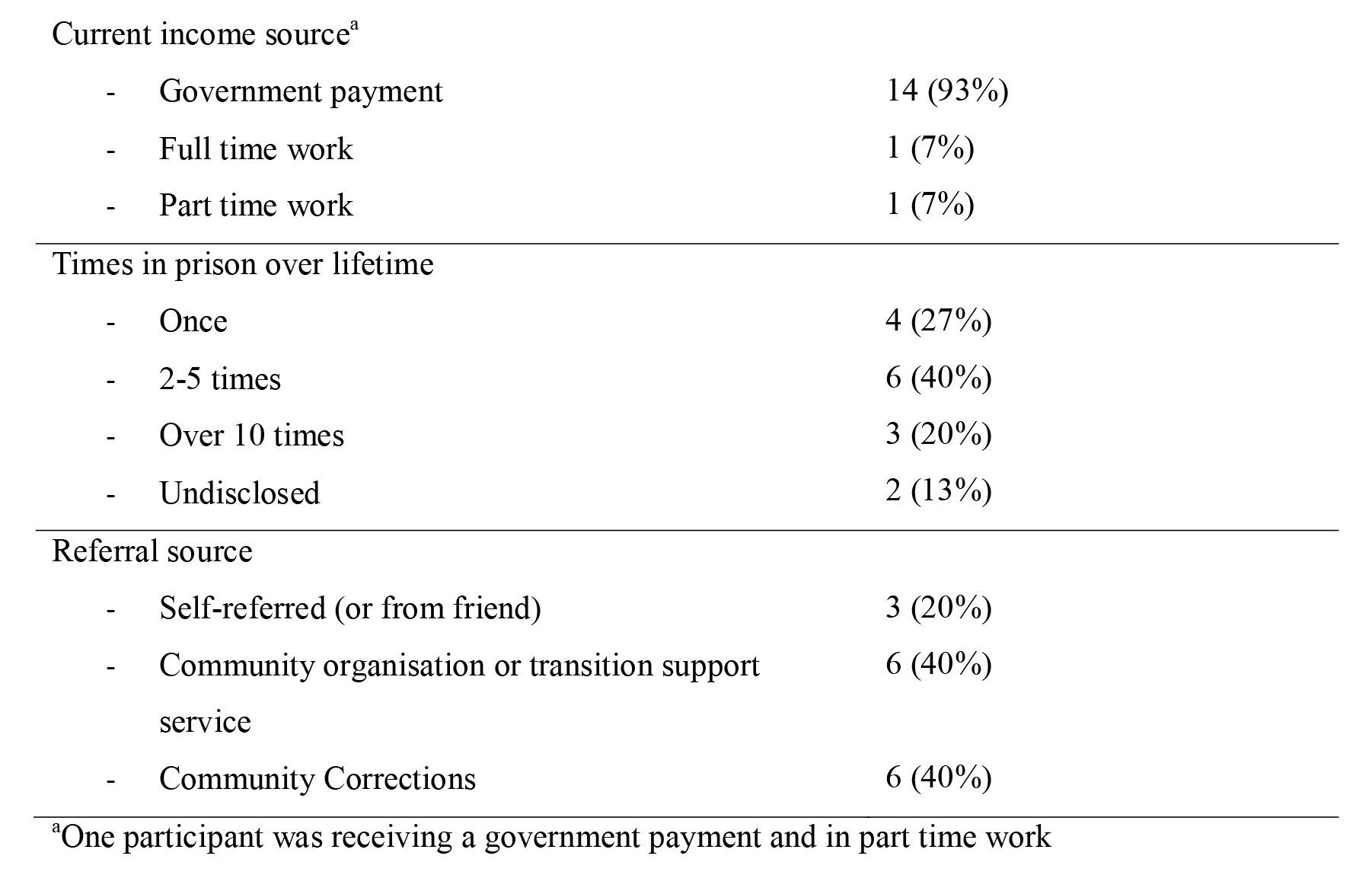
Demographic characteristics of interview participants (N=15)

| Demographic characteristic | *n* (%) |
| --- | --- |
| Mean Age (SD; Range) | 57 (6.3; 47-69) |
| Male | 14 (93%) |
| Aboriginal | 6 (40%) |
| Born in Australia | 15 (100%) |
| Mean months since leaving prison (Median, Range) | 8 (6, 1-24) |
| Most recent state of imprisonment |  |
| - New South Wales | 12 (80%) |
| - Victoria | 3 (20%) |
| Highest education |  |
| - No qualification | 4 (27%) |
| - Year 10 | 7 (47%) |
| - Diploma | 1 (7%) |
| - TAFE or trade certificate | 2 (13%) |
| - Postgraduate degree | 1 (7%) |
| Living Situation |  |
| - Own home or with family | 7 (47%) |
| - Public housing | 6 (40%) |
| - Renting privately | 2 (13%) |
| Current income source<sup>a</sup> |  |
| - Government payment | 14 (93%) |
| - Full time work | 1 (7%) |
| - Part time work | 1 (7%) |
| Times in prison over lifetime |  |
| - Once | 4 (27%) |
| - 2-5 times | 6 (40%) |
| - Over 10 times | 3 (20%) |
| - Undisclosed | 2 (13%) |
| Referral source |  |
| - Self-referred (or from friend) | 3 (20%) |
| - Community organisation or transition support service | 6 (40%) |
| - Community Corrections | 6 (40%) |

<sup>a</sup>One participant was receiving a government payment and in part time work

### Themes

The analyses resulted in six themes, organized around two global themes: ‘Release into a digital world’ and ‘Conditions for improvement’ (Figure 1). Whilst each theme is distinct, they also share associations (dashed lines).

**Figure 1.**
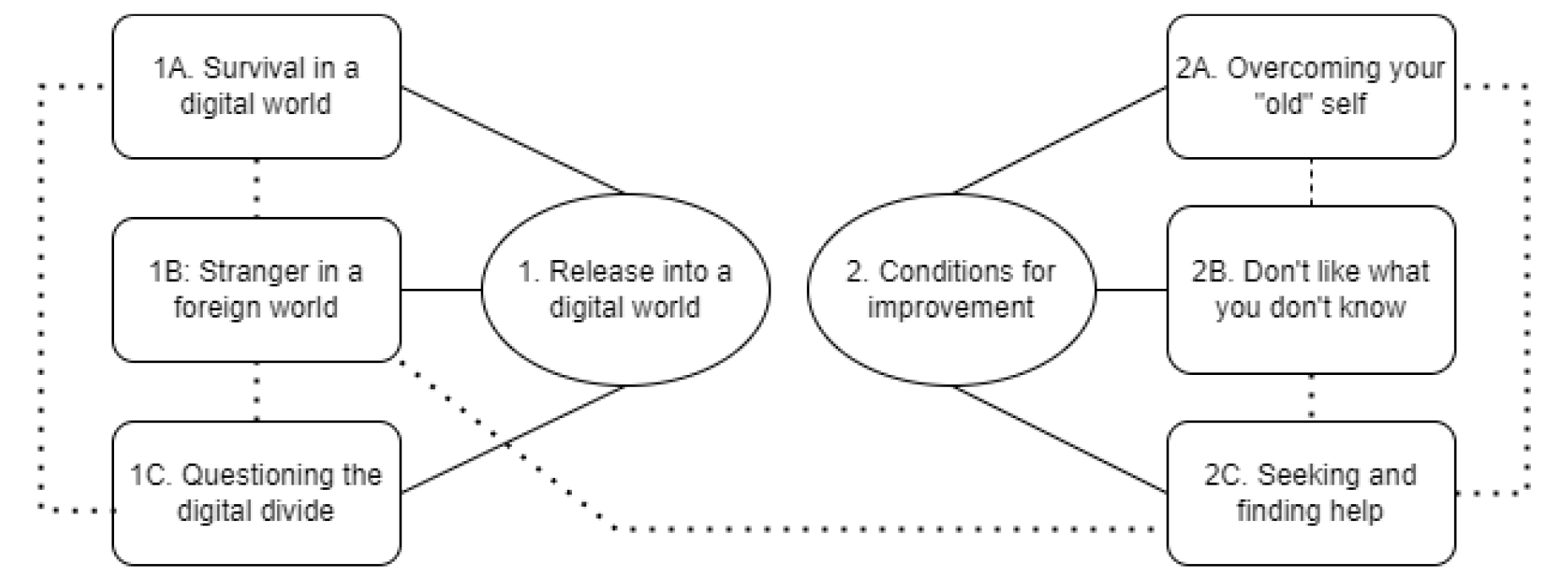
Global themes and themes resulting from reflexive thematic analysis of participant interviews.

### Global Theme 1: Release into a digital world

The first three themes depicted a confronting experience as participants left their technology-restricted prison environment to face a highly digitalized world. It included the associated struggle to survive and find a place in such a world, which they believed could have been mitigated with appropriate preparation in prison.

### Theme 1A: Survival in a digital world

Participants described post-release life as if entering a ‘digital world’. The frequent use of the terms “world” and “everything” in describing their experiences reflected the ubiquitous and widespread presence of technology. The way participants described this environment implied that learning to use technology was not a choice, but a necessity.

> *“It’s the new world now, you’ve got to get into using the technology.” P4*

> *“You’re in this world where everything, even to get a bus these days there’s some kind of technology because the ticket machines are all operated by little computers.” P13*

#### Given participants’ impression of such a world, it was unsurprising that digital skills were equated with survival or life skills

> *“Yes. I’m bloody hopeless at using tablets and computers and stuff. But I know now, I’ve got to learn, because, if I’m going to survive in this world.” P4*

> *“Some of us get out and we do not know how to do any of it. You need to be taught these skills, they’re life skills.” P2*

In describing the importance of digital literacy, participants often used examples of tasks that are crucial for resumption and re-establishment of a well-integrated life after prison. This most often included accessing government services for financial help, seeking employment, banking, and fulfilling parole requirements.

> *“When I came out, [my] bank account had been closed. Then you had to go and get your licence renewed and things like that you know? A lot of it’s on the computer now, or on your phone.” P11*

> *“Making sure that I complied with the parole conditions was pretty tough… having to use technology to do what they wanted me to do by way of reporting and various other things.” P3*

There was an immediacy in the need to use technology after being released. Acquiring a phone and learning to use it was often one of the first things participants did or planned to do upon release, and there was pressure to learn things quickly.

> *“The first thing I had to do [after release] was to get a mobile phone, and get the mobile phone set up.” P3*

> *“It was a three-hour drive from the prison to home, and that was basically me, learning whatever I could about the phone and how to how it functions.” P10*

### Theme 1B: Stranger in a foreign world

This theme captured feelings of estrangement and falling behind in this digital world. The idea of a digital divide was evident, and prison and prisoners were described to be alienated from the rest of the society as a result. First, a lack of digital literacy invoked feelings of being different to others. It was also a potential trigger for anxiety and stigma, by posing the risk of inadvertently revealing their ex-prisoner status.

> *“I think the internet has pretty much become a part of everyone’s day to day life, except for a prisoner.” P3*

> *"Not knowing this stuff, you stick out like a sore thumb. And anything that sort of has your head pop up outside of the norm really triggers anxiety” P6*

> *“It’s hard to fit in, because… I’ll give you an example. I was participating in [a community event]. When you finish, you have a [QR code]… I didn’t know how to bring that up on my phone. Because I didn’t even know you could. So, you get people looking like “What do you mean you don’t know?” Because everyone knows. And so, you go, “Oh, no, they know I’ve been in jail”.” P10*

The fast pace of technological advancement occurring in society rendered existing skills and resources irrelevant by the time people were released. This issue was pronounced for older people, who tend to have served longer sentences. Also, they were disadvantaged compared to younger prison leavers who had experience being part of a more digital world.

> *"Especially for anybody who’s done, say more than five to seven years, because things change so quickly, they don’t know what the world looks like. After 22 years… what’s a mobile phone, what’s any of those sorts of things?” P10*

> *“I think for younger people they were, I think, a little bit advantaged, so to speak, when they were leaving prison, because they were pretty much surrounded by a generation that was in tune with technology.” P3*

#### This pushed the ‘starting block’ for reintegration further back, and participants experienced the disadvantages of this lag both emotionally and functionally

> *“There’s nothing worse than technology moves on and on and the world leaves you behind.” P5*

> *“With technology the way it is, it has impacted me a fair bit, because I need to find out information. I’ve got to ask for help to do it. If I want information, say tonight, and I can’t get it. I gotta wait until tomorrow.” P4*

### Theme 1C: Questioning the digital divide

In this theme, participants considered the digital divide in prison as something that could have been mitigated. There was a tension between participants understanding the security issues that prevent technology in prison, but at the same time feeling that opportunities in prison were not well used. Key concepts in this theme included prison courses being outdated, irrelevant for reintegration, and underutilization of existing technology in prison.

First, it was common for participants to speak of having completed computer or IT courses in prison, but often this was a long time ago when computers were first emerging. In line with Theme 1B, this rendered their skills outdated after release.

> *“Years ago, this was back in like 1999/2000… I did all these computer courses there. Went through spreadsheets, you know, like payroll, you know, how to use your typing skills.”. P12*

> *“Yeah well I learnt, it was an old computer with a keyboard… in the 90s. I was I was okay… But now with new ones, I can turn it on but I’m no good.” P6*

Any courses they did receive addressed basic computer skills with limited application to real life. Participants believed that prison systems were deliberately resisting teaching internet use and tended to focus efforts on correcting offending behaviour. As in Theme 1A, digital literacy was equated to life skills.

> *“I can understand why they would want to block the internet [in prison]. But nor do they educate people how to use the internet for when they leave prison. What they don’t go into is the advancements in technology over the last few years.” P3*

> *“All the courses that they have for you to do are all to do with your offending, like drug courses, alcohol courses” P15*

Importantly, there has been increased access to tablets in prisons but a lack of support in learning to use them. This meant older people who often do not know how to use touch screen devices did not use them or needed to rely on younger inmates to help them, creating further digital division even within the prison.

> *“In most prisons, now, they have, like iPads. But no, they don’t teach you how to use it. They just expect you to know how to use it… Most young ones know how to use them. [those who don’t know how] just put them on the shelf and don’t worry about using them.” P5*

Participants argued for more opportunities in prison to learn digital literacy skills relevant to post-release life. In making suggestions for what this would look like, participants referred most to hands on practice or simulations of real-life situations. This resonates with Theme 1A and 1B in easing the friction as people enter a foreign world and addresses the immediate need for these skills after release.

> *“We could use augmented reality in prisons, because it’s not a security risk…. to simulate all these things that people may need to experience and learn how to do in the real world. And simple things as in how you use public transport, how to use tap and go at a supermarket.” P10*

> *“If they could somehow incorporate it into the prisons where they actually showed them how to use them and how to download an app and how to use the basic apps, teach them how to download it and how to use it, and they can actually have a go doing it themselves. It’s very hard to learn things, if ’you’re just, you know, are told, just download an app, and ’you’ll be right’. But the most people, particularly people over 50, 55, they’re not right, they can’t do it.” P1*

### Conditions for improvement

The remaining three themes centered on pertinent factors in an older prison leaver’s ability or willingness to improve their digital literacy.

### Theme 2A: Overcoming your “old” self

Participants struggled with the challenges of learning as an older person, as well as letting go of previous ways of life. Despite being older and thus more experienced with life in general, the “digital world” was one where things were reversed, and younger people were at an advantage. An intentional and effortful process of accepting this new order, and assuming a learning mindset in later life was needed.

> *“You get people that talk to you like you’re, that you’re dumb and stupid, because you’re 60 and they think you know everything. And I’m still learning about life and I’m still learning how to work things… because we’ve virtually got to start over from scratch, but at 60.” P4*

In addition to this learning mindset, it was apparent that participants would need to overcome ‘older’ ways of life. Participants had gotten used to a certain way of doing things over their lifetimes, and thus displayed resistance to newer, digital ways of doing these things.

> *“When I was at school, we didn’t have bloody computers like as far as I’m concerned you can do that with the old pen and paper you know? And I was good with pen and paper.” P11*

> *“My jobs were always outside jobs. I was a traffic worker, a forklift driver. I never, you don’t have a computer sitting on one of them.” P14*

### Theme 2B: Don’t like what you don’t know

This theme presents the notion of an emotional resistance that is often rooted in a lack of knowledge and confidence about technology, as well as their capacity for learning. Attitudes, feelings, and beliefs about technology, and about their own abilities to learn, were often interrelated and together determined a person’s openness to using technology and improving.

First, technology created an emotional response in people, the most common being fear and distrust. Importantly, these feelings stemmed from a lack of knowledge or understanding of technology.

> *“I’m not going to do online banking because I push the wrong button and I’m not putting an extra nought on there or something! I’m too scared. I’m scared of computers if that make sense?” P14*

> *“And then figuring out why they wanted to know, my email, you know, those sorts of things. Like who am I giving my email to, what do they need to know. And then just the linkages between different apps and why I needed to allow certain things to happen. What were the consequences if I did that?” P10*

A common belief that affected participants’ technology literacy was related to their intelligence or education level. When asked about their ability to use technology, participants often chose to speak about their intelligence, literacy, and schooling.

> *“Your IQ’s gotta be suitable for that [using the internet]. There’s so much I can’t do because of my literacy” P8*

This association went in both directions, where participants with high levels of education believed this was the reason for their proficiency, and vice versa.

> *“It’ll take a long time for me to work things out. Because I’ve got a thick head, and it takes a lot to go into this thick head of mine. Because I’ve only done six months of school all my life”. P4*

> *“So I can’t say that I’m naive about a lot of these things. Because fortunately, my mom and dad gave me a good education.” P3*

### Theme 2C: Seeking and finding help nearby

Finally, participants gained support for their digital literacy via ‘seeking’, or ‘finding’ it. First, seeking referred to directly asking for help or assistance with technology. Importantly, help was sought primarily from those in close physical proximity, such as members of their household and neighbours. For those who did not live with family, help was sought from support caseworkers and professional staff at telecommunications shops, and this was preferred to reaching out to families and friends.

> *“My neighbour he’s really good with computers and stuff. When I bought this phone I went over there and in 10 minutes bingo, everything was on my phone” P7*

> *“Yeah, I went to a phone guy downtown here, he’s helped a heap of ways.” P12*

> *“My daughter, sister and [caseworker]. They’re the three people that really I feel I can talk to but [caseworker] more so.” P8*

Participants tended not to seek help from friends or family who were not in close proximity, but did reach out to them when they were close by.

> *“Because I was down in Melbourne me mates pretty good on technology. When I was down in Melbourne I used it as much as possible, learned from friends down there.” P14*

There was a sense of burden implied in reaching out to others for help specifically regarding technology, and seeking help was described in terms such as ‘leaning on’ others and using their ‘precious time’.

> *“So I had to do I guess lean on the people that help me to get the phone set up with all the apps and stuff that I need.” P3*

> *“It took a lot of people, a lot of their precious time helping me. The [transition support service] I like when I first got out, it was a lot of their staff.” P6*

Those with help nearby thus counted themselves ‘lucky’. Whilst conversely those who were isolated, which older people were particularly at risk, would find it hard to seek help for their technology use.

> *“I can, use to get, you know, a friend or, or my daughters to help me out… I’m lucky I have a pretty good friends set up around [suburb] you know. I think I’m a lot more lucky than a lot.” P12*

> *“When you’re young, you got all your mates, you’ve got friends, you’ve got all.. help and everything, you know what I mean? But as you get older, everybody starts to dissipate and do their own thing and stuff like that, you know. You’re left basically fending for yourself. And if you haven’t learned that technology skill yet, you’re stuffed really.” P2*

Relatedly, many seemed to ‘find’ help via inadvertent exposure or proximity to others. This was particularly evident in participants who were living with other family members, which increased both access to a range of digital tools and support when needed.

> *“My mother and her husband are managers in their field sort of thing so they’ve got laptops and desktops and all that. If I need to send the resume, I get them to do it on their desktop computer. They put on the phone for me, yeah, that’s an app. I can operate the stereo turn it up and down off me phone.” P9*

In terms of improving their digital literacy, participants desired learning and support environments with face-to-face interaction alongside others who have experienced incarceration. Alongside several other themes this points to the pertinence of social and self-esteem issues related to stigma, learning at a pace and level that is adjusted to your ability, and the challenge of seeking help and re-learning in older age.

> *“Some kind of men’s groups where they can interact with other guys. And you, where it’s not embarrassing if they don’t know something.” P7*

> *“Everyone learns differently. At my own pace.” P13*

> *“I mean the library here, for example, runs a basic computer course, right? And I’ve noticed, there are a few older people attending that. So there would be nothing to stop a prisoner, ex-prisoner, joining in that group… but people say to you, you know, they want to get to know you or talk about who you are and where you’ve been and what you’ve done and all the rest of it. You compromise because you want to get to know these people, but you don’t want them to know you’ve been to jail. So I think a lot of ex-prisoners shy away from doing these community type education stuff. Because they’re paranoid, basically. P3*

## Discussion

### Key findings and implications

This study is the first to demonstrate the digital inequity and its effects experienced by older people after release from prison. Digital inequity arising from imprisonment in older age is perpetuated by a lack of support for digital literacy after release, with significant effects on the individual’s self-efficacy and identity, as well as their ability to adjust to social and economic functions of post-prison life and adhere to parole requirements. The study supports past findings that argue that technology adoption is a “sociotechnical process” that involves interwoven aspects of agency, structure and context surrounding the individual. Fang (2019) also drew attention to understanding social and structural factors in understanding the digital divide, and reports factors such as education, income, gender and generational status –collectively referred to as ‘privilege’- as important in determining digital inequities. These factors were found to be relevant to this population, who tend to have lower education, socioeconomic backgrounds and intergenerational trauma compared to those from the general population (Australian Institute of Health and Welfare, 2019; Honorato et al., 2016; Prost et al., 2021).

The findings were broadly aligned with the concerns of existing literature regarding the digital divide in prisons (Järveläinen & Rantanen, 2021; Jewkes & Reisdorf, 2016), whilst providing novel findings regarding the impact of this on older people during the post-release period. The digital divide is ‘deeper’ for older people leaving prison, who are prone to further inequity even in comparison to their younger imprisoned counterparts. Longer sentences that often go hand in hand with older age at release means prisoners miss out on more extended periods of technological advancement, making their life skills more outdated or irrelevant on release. This likely interacts with the accumulation of disadvantage that we already know exists for those who have served long prison sentences, due to loss of social connections, lost education and employment opportunities (Rakes et al., 2018).

A mismatch between low digital literacy and a highly digital world makes the post-release world a challenging place to navigate in older age. Older people are less likely than younger people to have been exposed to technology over their lifetimes, and so adjusting to such a world required going against many practices and attitudes that have been ingrained over their lifetimes. The findings depicted the immense psychological and mental adjustments that are required to adapt and fit into this digital world, alongside time pressures to learn quickly ensure ‘survival’. A shift in thinking is required here in recognizing that for this older group, post-release is not re-adjusting to life before entering prison, but newly adapting to a largely foreign world.

Whilst the motivation for improving digital literacy is present, the opportunities to prepare older people for release via digital literacy improvement, at least in Australia, appear underutilized. This aligns with the consistent evidence regarding a general lack of release planning practices (Hagos et al., 2021). According to this study, this may be because digital literacy for day-to-day life tasks is not considered a ‘criminogenic’ issue, i.e. directly related to recidivism. Most technology education in prison has focused on earning degrees or basic computer tasks, and release planning based on more traditional criminogenic needs such as drug use. Whilst these are important, this study demonstrates that many criminogenic factors are inevitably impacted by digital illiteracy, including finding accommodation, financial resources, education/employment, and community integration through leisure.

Taking a desistance mindset, considering digital illiteracy a key criminogenic issue and broadening pre-release planning to include a consideration of digital readiness will be beneficial for tackling recidivism for this group. Relatedly, researchers and policymakers have increasingly recognized the utility of using digital tools for rehabilitation and treatment for prisoners both whilst in prison and after release, such as rehabilitative apps, or digital means of supervision (e.g., Hwang et al., 2021; Perdacher et al., 2022; Seo et al., 2021). Older people in prison must be taught to use such technology for these rehabilitation initiatives to be successful.

Digital literacy also appears to have strong effects on self-identity and stigma. Digital illiteracy caused a feeling of estrangement from the rest of the world after release from prison. Moreover, it was a risk factor for stigmatization via inadvertently exposing a person’s imprisonment history. Both are important factors implicated both in positive psychosocial adjustment in later life, and in desistance from crime. The need for digital literacy also amplified the social challenges associated with release from prison in old age. Those who had access to social resources were advantaged in improved access to technology as well as support to improve their literacy. This reinforced the need for social networks, which can often be lost in this group due to long sentences and more serious crimes.

### Future work

Targeted digital literacy initiatives are needed for older prison leavers, and these should be delivered both in prison and in the community. Whilst solutions will be local and consider the resources and infrastructure available to each locale, broad reviews of existing evidence and examples of best practice will be important to ensure best allocation of resources. This should begin with a review of digital literacy programs already available for older adults in the community, for example via public spaces such as libraries, which have been recognized as good potential spaces for tackling digital inequity in the community (Strover, 2019). Then, the applicability of these courses to the unique needs of this population should be considered.

Also useful will be a similar review of current practices in technology provision and technology-related courses in correctional centers both nationally and internationally. Regarding this, researchers have highlighted the difficulty in locating publicly available information regarding internet and technology-related policies in correctional centers, at least in Australia (Kerr & Willis, 2018). It will be important for correctional centers to recognize the importance of the issue and the benefit in working together with researchers and community-based stakeholders to move towards evidence-based solutions.

Important considerations for such initiatives include the likelihood of low education and literacy levels, their often-transient living situations and personal financial constraints. Social issues relating to stigmatization of this group may also mean that programs specific to prison leavers may be preferable. However, other creative solutions where this group may learn alongside and form social connections with other members of the community whilst mitigating the risk of stigmatization would also be valuable. In any case, specific intervention development should occur in further consultation with this group and relevantly experienced stakeholders.

Another important consideration for designing literacy programs is the difficulty in help-seeking, and feelings of burdensomeness found in this group. This may reflect both a culture of ‘hardness’ in prisons, and the well-evidenced negative relationship between masculinity and help-seeking. Older prisoners are prone to social difficulties within prisons, which may further impact passivity and discourage self-efficacy. In one study in the US, it was found that improving digital literacy in prisoners had the effect of improving self-efficacy (Champion & Edgar, 2013). Therefore, we may expect that equipping prison leavers with digital literacy may also have positive effects on their self-efficacy and help-seeking.

The courses available in prisons should emulate real life situations, with a view to preparing the person for success after prison and reducing the ‘foreignness’ of post-release life identified in this study. In line with the conclusions of a recent study of Finnish prisoners, there is a particular need to teach prisoners to use digitized health care and social welfare services that they will need to access immediately after release (Rantanen et al., 2022). These should be delivered alongside initiatives to improve existing release planning practices that are broader in focus beyond basic criminogenic needs.

To fully understand the digital literacy level and effects on reintegration of prisoners, additional quantitative data should be collected using validated and appropriate measures of digital literacy. For example, a recent collaborative effort between universities and government agencies in Australia has seen the development of the Australian Digital Inclusion Index (Thomas et al., 2021) which would provide a useful means of examining this group specifically in light of the rest of community.

Economic analyses that highlight the benefits and costs associated with investing in digital literacy for this group would be particularly useful for decisionmakers.

### Limitations

The study included several limitations which should be considered when interpreting the findings. First, despite purposive sampling efforts for many months, the researchers were unable to recruit more than one female to the study. Whilst females represent a minority of prison leavers (6% of older prisoners in Australia), they have high and unique needs that differ to that of males (Barry et al., 2020; Handtke et al., 2015). It will be important for a future dedicated study of older female prison leavers to be conducted.

The sample was also all sourced from two of seven Australian jurisdictions (New South Wales and Victoria). Each jurisdiction and correctional center have differing practices both related to in-prison technology delivery and release. The findings of the study that are related to specific practices should thus be interpreted with caution. Given this, New South Wales and Victoria account for over 50% of Australia’s prisoner population. Also, whilst not explicitly collected and reported in the study, when speaking about their histories of imprisonment, many participants reported having been incarcerated across different states outside of New South Wales and Victoria. Nonetheless, the trend towards accepting and integrating technology into prison practices and the challenges of release into a technology-driven modern society, are universal.

The recruited sample was relatively ‘young’ compared to other studies of older people, with the oldest participant still being under 70 years of age. Those who are older may have unique and heightened challenges stemming from the ‘accelerated ageing’ that this group tend to exhibit (Greene et al., 2018). These findings are nonetheless in line with that reported by stakeholders and staff with professional experience supporting older prison leavers in another study by the research team (Hwang et al, preprint).

Finally, as with all research with justice-involved populations, there was a risk of responder bias (e.g., coercion, social desirability) due to power imbalances between the research staff and the participant. This was particularly relevant to the study recruitment pathway that involved corrective services staff who were currently supervising some of the participants (40% of the sample). The risk of coercion to participate in the study was dealt with in a way that appeased multiple ethical review committees. Staff at corrective services or transition support services were only involved in first offering the study to participants, after which all contact was made only between the participant and research team. Anonymity of responses and the independence of the research from these organisations was made clear throughout the recruitment and interview process. We did not detect any findings that may have resulted from social desirability.

## Data Availability

The authors do not have permission to share data.

## Acknowledgements

The authors wish to thank ethics for providing financial support for the conduct of this research study. The authors also wish to acknowledge the support from the NSW Community Restorative Centre.

## Notes

### Competing Interest Statement

The authors have declared no competing interest.

### Funding Statement

This study was funded by the Australian Association of Gerontology Hal Kendig Research Development Program

